# High-Fidelity Synthetic Data Replicates Clinical Prediction Performance in a Million-Patient Diabetes Cohort

**DOI:** 10.1101/2025.07.20.25331852

**Authors:** Víctor M. de la Oliva-Roque, David P. Kreil, Joaquín Dopazo, Francisco Ortuño, Carlos Loucera

## Abstract

Synthetic data generated using generative models trained on real clinical data offers a promising solution to privacy concerns in health research. However, many efforts are limited by small or demographically narrow training datasets, reducing the generalizability of the synthetic data. To address this, we used real-world clinical data from nearly one million individuals with diabetes in the Andalusian Population Health Database (BPS) to generate a comprehensive longitudinal synthetic dataset.

We employed a dual adversarial autoencoder to produce synthetic data and evaluated its utility in a clinical machine learning (ML) task: predicting the onset of chronic kidney disease, a common diabetes complication. Models trained on synthetic data were assessed for their ability to reproduce patterns and predictive behaviors observed in real data. Performance and stability were compared across models trained on real, synthetic, and hybrid datasets. Models trained exclusively on synthetic data achieved AUROC scores comparable to real-data models (0.70 vs. 0.73) and showed high stability in feature importance rankings (weighted Kendall’s τ > 0.9). Notably, combining synthetic and real data did not improve performance.

Our findings demonstrate that high-fidelity synthetic longitudinal data can replicate real data performance in clinical ML, supporting its use in research while preserving patient privacy. This represents a significant step toward more collaborative and privacy-preserving healthcare data ecosystems.

## 1 Introduction

In healthcare settings, real-world data (RWD) refers to observational data that is not collected as part of a defined clinical trial or observational study, but that are rather generated from real-world settings such as patient electronic health records (EHRs) (1). As such, this data is not constrained by the inclusion and exclusion criteria intrinsic to the aforementioned studies (2), allowing them to capture the heterogeneity within a patient population and providing a more comprehensive picture of clinical practice and outcomes (1, 3). Furthermore, as these data can comprise any given patient’s longitudinal clinical history, they can be invaluable in observing patient’s disease trajectories, understood as the sequential and directional co-occurrence of diseases (4). Indeed, various disease trajectories have been observed from population-wide registry data (5) and differing trajectories have proven to have an impact on a patient’s healthcare burden (6). Moreover, leveraging longitudinal information has shown to outperform static information in prediction studies (7, 8).

Nevertheless, given the highly sensitive nature of RWD, which can contain information on patient demography, clinical health and even financial situation, its access is subject to stringent data protection regulations such as the General Data Protection Regulation (GDPR) (3). While various methods have been proposed to anonymize this identifiable patient data, re-identification is still a risk (9) and achieving a balance between the degree of data anonymization and usability remains a challenge for widespread data sharing (10).

As such, synthetic data generation through generative artificial intelligence (AI) models has been proposed as a method to circumvent these problems. This approach can furthermore be used for data augmentation, increasing dataset size, and improving diversity within datasets (11). Namely, Generative Adversarial Network (GAN) (12) and Variational Autoencoder (VAE) (13) architectures have been successful in synthesizing EHR data for various clinical purposes. More recently, Dual Adversarial Autoencoders (DAAE) have been used to synthesize sequences of set-valued medical records (14), addressing the longitudinal nature of EHRs in order to recreate disease trajectories.

This implementation, however, has been carried out on the MIMIC database, which is a single-center database containing data on patients admitted to critical care units (15). This data is limited in its scope, as patients admitted to critical care units are a relatively small and focused population subject to constant and continuous monitoring. Furthermore, this data is focused on patient’s critical care treatment and does not span their previous clinical history. As such, this does not address many of the challenges inherent to RWD and consequently in generating synthetic data from them. Namely, the fact that they tend to be poorly structured, incomplete, heterogeneous, and prone to measurement errors and biases (3). On the other hand, this implementation misses the various benefits of RWD obtained from large clinical databases, namely the long spanning and diverse patient-level longitudinal data available for varying cohorts, which can provide a more comprehensive picture of real outcomes and trajectories.

Given these limitations, we have implemented a DAAE architecture to generate high-fidelity synthetic data using data extracted from the Andalusian Population Health Database (BPS, due to its acronym in Spanish) (16). This database contains longitudinal information derived from 15 million patient’s EHRs from the Andalusian public health system. Specifically, we have used 1 million EHRs from diabetic patients in order to generate synthetic patient disease trajectories. We measure the utility of the generated trajectories through train on synthetic, test on real (TSTR) performance (17). Namely, we predicted a given end-point by training on a synthetic dataset and evaluating on the real test set, comparing this performance to that of a model trained with real data. We furthermore compared feature rankings between models trained with synthetic and real data. We then tested the potential of our generated data for data augmentation by training a hybrid model with both real and synthetic data, and evaluate its performance.

In particular, we have predicted the onset of chronic kidney disease (CKD), a common complication in diabetic patients, using their prior comorbidities. Type 2 diabetes, particularly, has been associated with a vast spectrum of CKD etiologies, with various studies finding its onset to be compounded due to the patient’s medical history (18).

## 2 Materials and Methods

### 2.1 Data

The Andalusian Biomedical Research Ethics Coordinating Committee approved the study entitled: “Generation of synthetic patients for the study of diseases and testing of algorithms for health research” (September 24, 2019, Acta 08/19) and waived informed consent for the secondary use of clinical data for research purposes.

Longitudinal real-world data were extracted from the BPS (16), based on EHRs from all users of the Andalusian public health system. A retrospective cohort of 1,062,633 individuals were obtained from the BPS with a registered diabetes mellitus diagnosis in Andalusia from 2003 to 2022. After filtering for valid gender coding and adult-onset diabetes diagnoses (age≥18) a total of 908,673 remained. As type-I diabetes mellitus (ICD10 E10) is rarely diagnosed at this age, the vast majority of our cohort consists of type-II diabetes patients (ICD10 E11). The average age at diagnosis was 60.6±13.83 years. Comorbidities without visit dates were excluded. The final dataset included 5,869,024 pathology diagnoses across 4,875,272 medical visits from 1930 to 2022. For end-point prediction, patients with any prior diabetes diagnosis before 2003 and patients with a CKD diagnosis before their diabetes diagnosis date were removed, resulting in a final cohort of 766,904 patients. Of these, 78,729 (10.27%) had a CKD diagnosis after their diabetes diagnosis. Data also included 79 additional comorbidities. Diagnoses covered both prior history and follow-up, up to 2022. Patients with a CKD diagnosis were those with any of the ICD10 codes in Table 1.

**Table 1.** ICD10 codes used for CKD patient identification.

| ICD10 | ICD10 description |
| --- | --- |
| N18 | Chronic kidney disease (CKD) |
| D63.1 | Anemia in chronic kidney disease |
| E08.2 | Diabetes mellitus due to underlying condition with kidney complications |
| I13 | Hypertensive heart and chronic kidney disease |
| E11.2 | Type 2 diabetes mellitus with kidney complications |
| E10.2 | Type 1 diabetes mellitus with kidney complications |
| I12 | Hypertensive chronic kidney disease |
| E09.2 | Drug or chemical induced diabetes mellitus with kidney complications |
| E13.2 | Other specified diabetes mellitus with kidney complications |

Patients were encoded as sequences of health system visits, represented by age at visit and diagnoses. Each patient’s record includes sex and an ordered list of diagnosis events, one being diabetes.

### 2.2 Secure data management

The data management circuit was designed to minimize the relative risk as described in the Impact Assessment on Data Protection analysis (19) and following the regulation for the use of medical data for research purposes in Andalusia (20). Briefly, the data corresponding to the study approved by the ethics committee is requested to BPS. There, the data is extracted and pseudonymized by BPS personnel and then transferred to the Platform for Generation of Medical Evidence (PAGEM) (21), a Secure Processing Environment (SPE) specifically designed for the secure analysis of data protected by the GDPR, being compliant with the definition of SPE as described in Article 50 of the Resolution of the European Parliament of 24 April 2024 on the proposal for a Regulation of the European Parliament and of the Council on the EHDS (22).

### 2.3 Generative Adversarial Training

Data were split into a training (80%) and leave-out test set (20%) and synthetic data was generated exclusively with the train set. Dual adversarial autoencoders (DAAE) were used to learn realistic sequences from medical data by jointly modeling latent and discrete data distributions (14). The model was configured with a batch size of 256 and trained for 500 epochs on an NVIDIA Tesla V100 GPU with 32GB.

### 2.4 End-point prediction

Patients with a chronic kidney disease diagnosis before their diabetes diagnosis were removed from the at-risk population for end-point prediction in both the real and synthetic datasets.

We trained an Explainable Boosting Machine (EBM) (23) with default hyperparameters and no feature interactions using the aforementioned training set used for synthetic data generation, the generated synthetic dataset, as well as a model trained using a mix of both real and synthetic data.

Training features were generated by converting patient comorbidities diagnosed before their end-point diagnosis to binary variables. As controls did not experience the end-point, they were randomly matched with a case end-point date posterior to their diabetes diagnosis date. Finally, ten repeated stratified 10-fold splits of the training set were generated and separate models trained on each fold. We evaluated the difference in prediction probabilities on the leave-out test set between these models trained on data splits and the model trained with the entire training set and report means and standard deviations. Furthermore, we calculated the Kendall’s weighted τ on feature rankings both within and between training sets and report distributions, medians and inter-quartile ranges.

Predictions on the test set were carried out considering the comorbidities diagnosed before patient’s diabetes diagnosis date, thus simulating a patient’s evaluation of CKD risk at the time of diabetes diagnosis.

## 3 Results

### 3.1 Synthetic Generation

A synthetic cohort of 800,000 individuals was generated using DAA) (14), based on the real dataset. Quality checks excluded non-diabetic individuals, empty visits, redundant chronic disease entries, and a lack of associated patient sex. For age data, the highest value per visit was retained, and missing ages were interpolated from adjacent visits. The final synthetic cohort included 711,080 individuals for analysis, of which 49,796 (7.00%) had a CKD diagnosis. After removing individuals with end-points before their associated diabetes age, a total of 685,798 remained. Of these, 33,726 (4.92%) had an associated end-point after their diabetes diagnosis age.

### 3.2 End-point prediction performance

Both models trained on real and on synthetic data showed a steady increase in their predictive performance when evaluating patients whose diabetes diagnosis date was in more recent calendar years, until reaching relative stabilization in 2014. From this year onwards, models trained with real data showed an AUROC of 0.73 and models trained with synthetic data an AUROC of 0.70, representing a 4% decrease in AUROC. However, the hybrid model trained with both real and synthetic data showed no improvement compared to the model trained solely with real data (AUROC of 0.73) Figure 1A. Yearly mean AUROC with standard deviations, as well as total positive and negative class individuals are presented in Table 2.

**Table 2.** AUROC mean and standard deviations, as well as class counts by diabetes diagnosis year across Hybrid, Real, and Synthetic training sets.

| Diabetes year | Metric | Hybrid | Real | Synthetic |
| --- | --- | --- | --- | --- |
| 2003 | AUROC Mean (SD) | 0.62 (2.44e-04) | 0.62 (3.19e-04) | 0.62 (7.08e-04) |
|  | N negative class | 4544 | 4544 | 4544 |
|  | N positive class | 1474 | 1474 | 1474 |
| 2004 | AUROC Mean (SD) | 0.64 (2.31e-04) | 0.64 (3.41e-04) | 0.63 (5.60e-04) |
|  | N negative class | 5390 | 5390 | 5390 |
|  | N positive class | 1425 | 1425 | 1425 |
| 2005 | AUROC Mean (SD) | 0.65 (2.20e-04) | 0.65 (2.44e-04) | 0.64 (7.19e-04) |
|  | N negative class | 5166 | 5166 | 5166 |
|  | N positive class | 1330 | 1330 | 1330 |
| 2006 | AUROC Mean (SD) | 0.66 (2.60e-04) | 0.66 (3.06e-04) | 0.65 (7.80e-04) |
|  | N negative class | 5682 | 5682 | 5682 |
|  | N positive class | 1252 | 1252 | 1252 |
| 2007 | AUROC Mean (SD) | 0.67 (2.68e-04) | 0.67 (2.70e-04) | 0.66 (7.50e-04) |
|  | N negative class | 7016 | 7016 | 7016 |
|  | N positive class | 1460 | 1460 | 1460 |
| 2008 | AUROC Mean (SD) | 0.66 (3.10e-04) | 0.67 (3.29e-04) | 0.65 (9.60e-04) |
|  | N negative class | 7361 | 7361 | 7361 |
|  | N positive class | 1292 | 1292 | 1292 |
| 2009 | AUROC Mean (SD) | 0.69 (2.83e-04) | 0.69 (2.95e-04) | 0.68 (8.17e-04) |
|  | N negative class | 7414 | 7414 | 7414 |
|  | N positive class | 1136 | 1136 | 1136 |
| 2010 | AUROC Mean (SD) | 0.69 (2.98e-04) | 0.69 (4.00e-04) | 0.68 (8.66e-04) |
|  | N negative class | 7663 | 7663 | 7663 |
|  | N positive class | 1100 | 1100 | 1100 |
| 2011 | AUROC Mean (SD) | 0.70 (3.07e-04) | 0.70 (3.34e-04) | 0.68 (9.78e-04) |
|  | N negative class | 7444 | 7444 | 7444 |
|  | N positive class | 943 | 943 | 943 |
| 2012 | AUROC Mean (SD) | 0.71 (3.25e-04) | 0.71 (3.73e-04) | 0.69 (9.47e-04) |
|  | N negative class | 6730 | 6730 | 6730 |
|  | N positive class | 715 | 715 | 715 |
| 2013 | AUROC Mean (SD) | 0.74 (3.12e-04) | 0.73 (3.51e-04) | 0.71 (9.02e-04) |
|  | N negative class | 7103 | 7103 | 7103 |
|  | N positive class | 703 | 703 | 703 |
| 2014 | AUROC Mean (SD) | 0.74 (3.35e-04) | 0.74 (4.04e-04) | 0.71 (9.66e-04) |
|  | N negative class | 7011 | 7011 | 7011 |
|  | N positive class | 548 | 548 | 548 |
| 2015 | AUROC Mean (SD) | 0.74 (3.91e-04) | 0.74 (4.94e-04) | 0.71 (1.17e-03) |
|  | N negative class | 6457 | 6457 | 6457 |
|  | N positive class | 437 | 437 | 437 |
| 2016 | AUROC Mean (SD) | 0.75 (4.38e-04) | 0.76 (3.96e-04) | 0.70 (1.54e-03) |
|  | N negative class | 6561 | 6561 | 6561 |
|  | N positive class | 383 | 383 | 383 |
| 2017 | AUROC Mean (SD) | 0.73 (3.75e-04) | 0.73 (3.86e-04) | 0.70 (1.12e-03) |
|  | N negative class | 6820 | 6820 | 6820 |
|  | N positive class | 297 | 297 | 297 |
| 2018 | AUROC Mean (SD) | 0.74 (5.59e-04) | 0.75 (6.12e-04) | 0.70 (1.63e-03) |
|  | N negative class | 6951 | 6951 | 6951 |
|  | N positive class | 277 | 277 | 277 |
| 2019 | AUROC Mean (SD) | 0.71 (4.94e-04) | 0.72 (5.24e-04) | 0.68 (1.53e-03) |
|  | N negative class | 7318 | 7318 | 7318 |
|  | N positive class | 226 | 226 | 226 |
| 2020 | AUROC Mean (SD) | 0.76 (6.72e-04) | 0.77 (8.07e-04) | 0.72 (1.56e-03) |
|  | N negative class | 6464 | 6464 | 6464 |
|  | N positive class | 152 | 152 | 152 |
| 2021 | AUROC Mean (SD) | 0.68 (6.16e-04) | 0.69 (8.05e-04) | 0.67 (1.69e-03) |
|  | N negative class | 9712 | 9712 | 9712 |
|  | N positive class | 109 | 109 | 109 |
| 2022 | AUROC Mean (SD) | 0.72 (1.11e-03) | 0.71 (1.31e-03) | 0.67 (3.69e-03) |
|  | N negative class | 6511 | 6511 | 6511 |
|  | N positive class | 34 | 34 | 34 |

**Figure 1.**
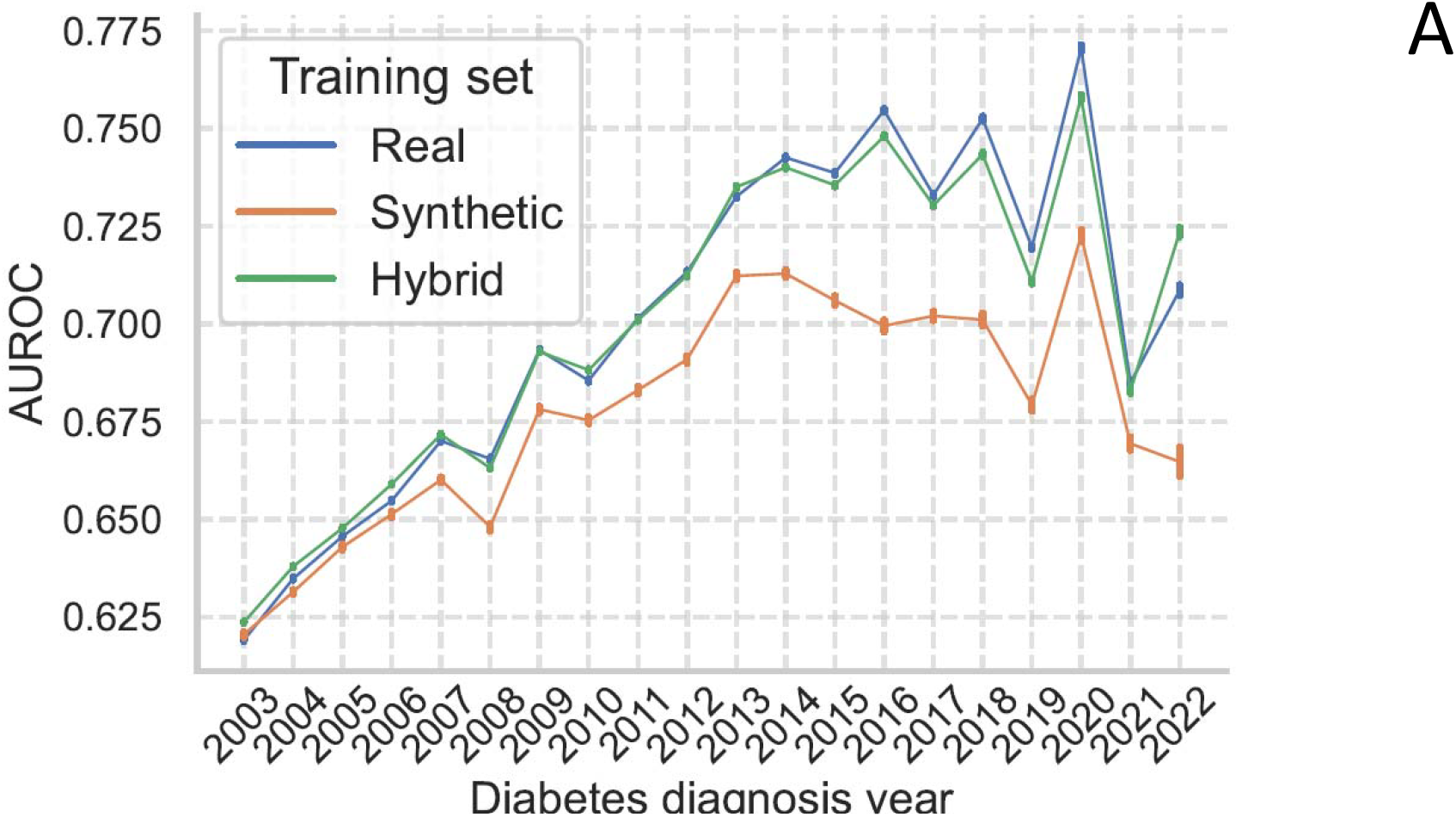
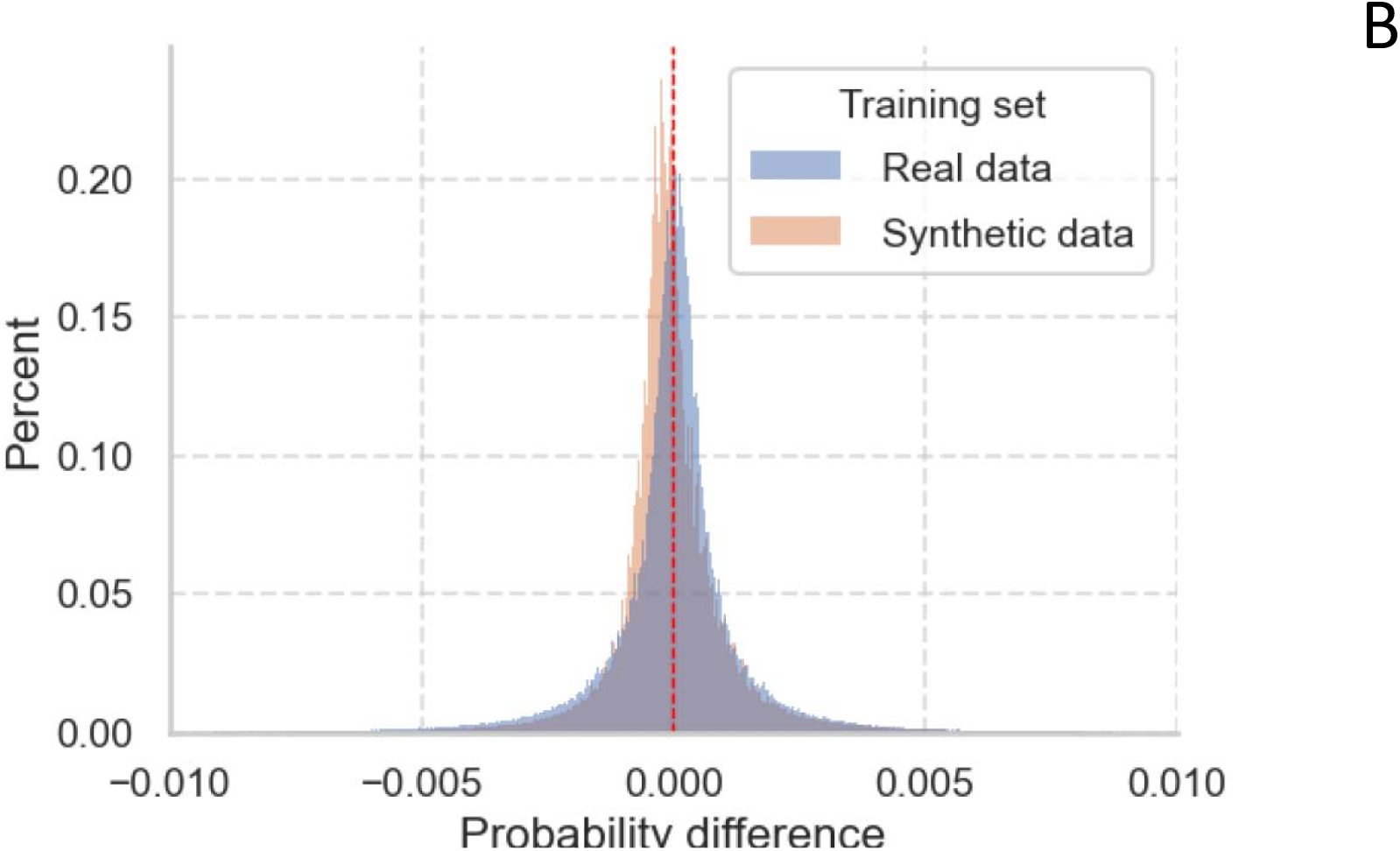
(A) Mean AUROC across years of models trained with dataset splits of real-only, synthetic-only and a hybrid of both. Error bars indicate standard deviations (B) differences in prediction probabilities on the test set between models trained with the full training set and splits (trimmed at x=-0.1 and x=0.1 for visualization).

Prediction probabilities between models trained with the entire training set and those trained on training set splits were small and heavily centered on 0 in both training sets Figure 1B. The median difference between these was namely 5.0×10^−5^ [−4.4×10^−4^, 5.0×10^−4^] for models trained on real data and −1.1×10^−4^ [−5.0 × 10^−4^, 3.9 × 10^−4^] for models trained on synthetic data.

Feature ranking stability, measured by Kendall’s hyperbolic-weighted τ, was high for models trained on splits of both real data (median τ = 0.966 [0.963, 0.969]) and synthetic data (median τ = 0.953 [0.948, 0.957]), demonstrating strong internal consistency (Figure 2A). Furthermore, the rankings learned from synthetic data showed good agreement with those from real data, yielding a median weighted τ of 0.742 [0.731, 0.754] (Figure 2B).

**Figure 2.**
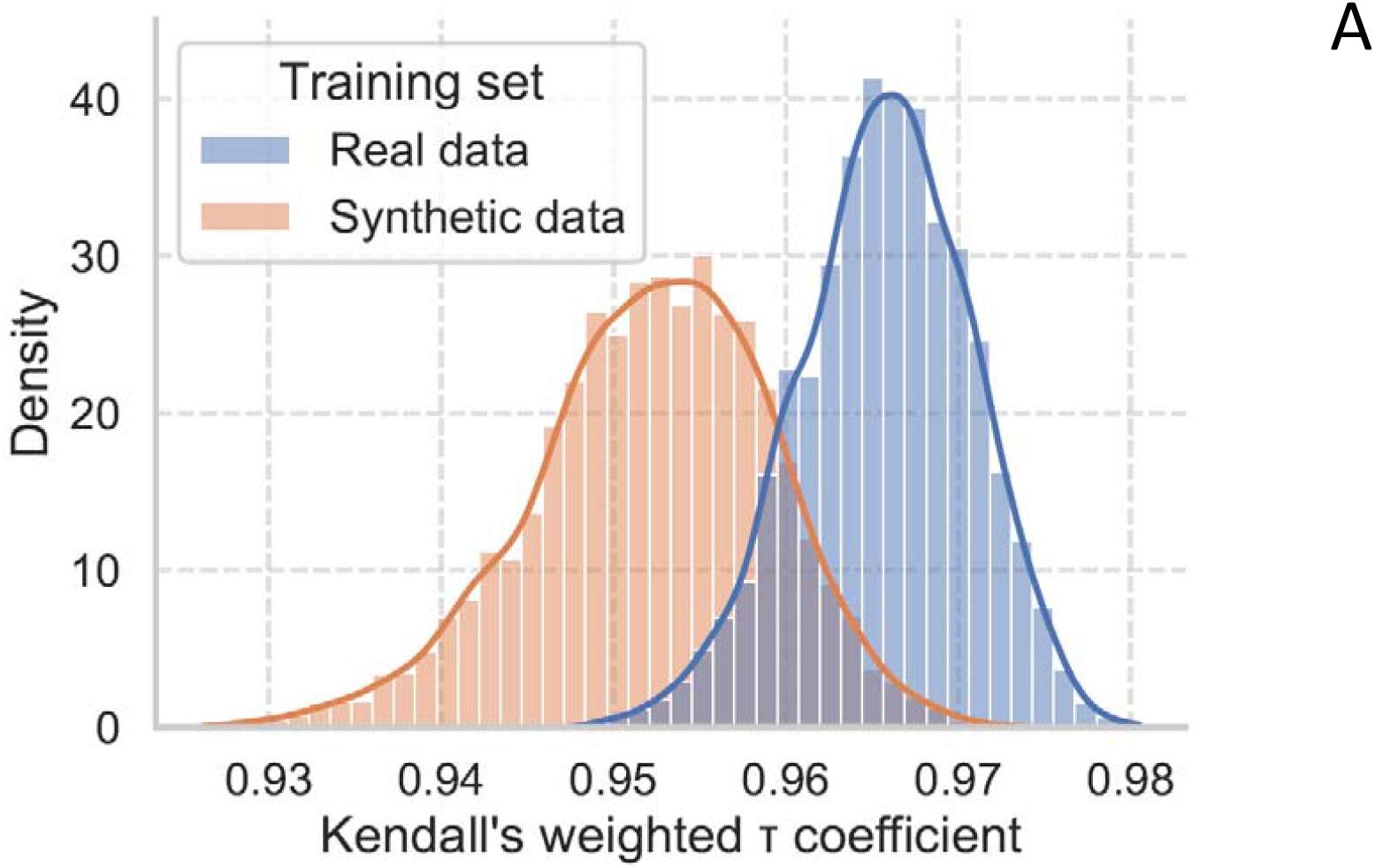
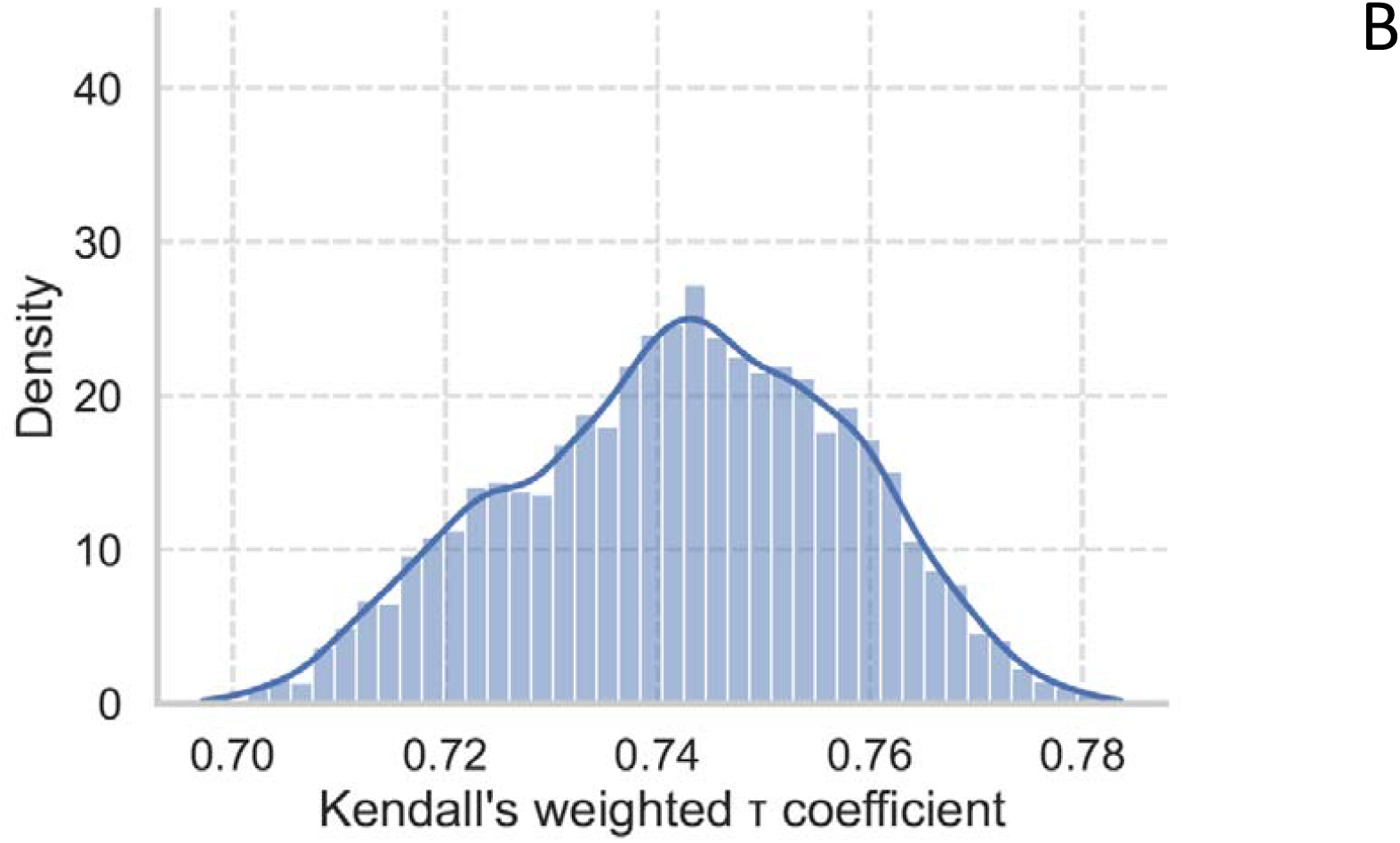
(A) Within training set Kendall’s weighted τ of feature rankings (B) Between training set Kendall’s weighted τ of feature rankings

## 4. Discussion and future work

### 4.1 Discussion

In this work, we have successfully generated high-fidelity longitudinal synthetic data from a large and diverse diabetic RWD cohort, and shown its utility in a relevant clinical scenario, namely the ML assessment of CKD risk in diabetic patients. This synthetic model shows a 4.11% decrease in performance as measured by AUROC relative to the model trained with real data when evaluating on calendar years after performance stabilization. In addition, both our synthetic and real models show good agreement between their feature rankings, indicating that high-relevance features for the specified end-point remain so in the synthetic dataset. This performance was furthermore highly consistent across dataset splits for both models.

This shows the potential of synthetic data generation not only in well-curated clinical databases, but also in RWD, not only tacking the challenges that this type of data presents, but also exploiting the richness that this data provides. Given this fact, this dataset and others like it can be used to study various other downstream tasks besides that presented here. As these datasets do not correspond to any real individuals, they can freely be made public for the wider research community, thus accelerating advancements in clinical research. This shows to be a promising solution to the challenges currently faced by researchers in accessing valuable clinical data.

### 4.2 Future work

In order to further test the generated dataset’s utility, our methodology should be applied to other end-points within the BPS. In the context of diabetic patients and their risks, retinopathy and/or heart failure are interesting candidates. Furthermore, in order to fully test and exploit the generated disease trajectories, deep learning architectures which can leverage time-series data can be applied for end-point prediction.

Furthermore, other generative models can be used to generate the synthetic data. Their quality and utility can then be compared to that achieved by the DAEE.

Finally, further endeavors at improving model performance through augmentation with synthetic data should be done. This could be done at the level of synthetic data quality improvement, or rather by exploiting more complex real datasets. Indeed, models trained on real data did not show significantly improvement when training on the entire dataset as opposed to dataset splits (as seen by the nearly identical prediction probabilities). This could indicate that the informativeness of the real dataset used is limited in predicting the end-point and therefor adding more data does not improve performance. In this regard, different data modalities such as blood analysis values, medication and/or procedures could be added to both synthetic data generation and end-point prediction.

## Data availability

The data used for the training of the generative model are available in the Population Health Database repository (16), subject to controlled access according to the regulation for the use of medical data for research in the Andalusian Health System (20).

The synthetic data produced in the present study are currently available after signing an agreement within the “Synthetic Clinical Health Records Challenge” of the CAMDA conference (https://www.camda.info/)

## Acknowledgments

This study was supported in part by grants from the Spanish Ministry of Science and Innovation (PID2023-152380OB-C21, PRTR-C17.I1), the Institute of Health Carlos III (PMP24/00024), the Consejería de Salud y Consumo from the Junta de Andalucía (EXC-2023-01, IE19_259 FPS, PAIDI 2020, IE19_259 FPS).

## References

1. Makady A, de Boer A, Hillege H, Klungel O, Goettsch W. What is real-world data? A review of definitions based on literature and stakeholder interviews. Value in health. 2017;20(7):858–65.

2. Kandi V, Vadakedath S. Clinical trials and clinical research: a comprehensive review. Cureus. 2023;15(2).

3. Liu F, Panagiotakos D. Real-world data: a brief review of the methods, applications, challenges and opportunities. BMC Medical Research Methodology. 2022;22(1):287.

4. Jørgensen IF, Haue AD, Placido D, Hjaltelin JX, Brunak S. Disease trajectories from healthcare data: methodologies, key results, and future perspectives. Annual review of biomedical data science. 2024;7(1):251–76.

5. Jensen AB, Moseley PL, Oprea TI, Ellesøe SG, Eriksson R, Schmock H, et al. Temporal disease trajectories condensed from population-wide registry data covering 6.2 million patients. Nature communications. 2014;5(1):1–10.

6. Beck MK, Westergaard D, Jensen AB, Groop L, Brunak S, editors. Temporal order of disease pairs affects subsequent disease trajectories: the case of diabetes and sleep apnea. PACIFIC SYMPOSIUM ON BIOCOMPUTING 2017; 2017: World Scientific.

7. Nielsen AB, Thorsen-Meyer H-C, Belling K, Nielsen AP, Thomas CE, Chmura PJ, et al. Survival prediction in intensive-care units based on aggregation of long-term disease history and acute physiology: a retrospective study of the Danish National Patient Registry and electronic patient records. The Lancet Digital Health. 2019;1(2):e78–e89.

8. Placido D, Yuan B, Hjaltelin JX, Zheng C, Haue AD, Chmura PJ, et al. A deep learning algorithm to predict risk of pancreatic cancer from disease trajectories. Nature medicine. 2023;29(5):1113–22.

9. Langarizadeh M, Orooji A, Sheikhtaheri A. Effectiveness of anonymization methods in preserving patients’ privacy: a systematic literature review. Health Informatics Meets eHealth. 2018:80–7.

10. Zuo Z, Watson M, Budgen D, Hall R, Kennelly C, Al Moubayed N. Data anonymization for pervasive health care: systematic literature mapping study. JMIR medical informatics. 2021;9(10):e29871.

11. van Breugel B, Liu T, Oglic D, van der Schaar M. Synthetic data in biomedicine via generative artificial intelligence. Nature Reviews Bioengineering. 2024;2(12):991–1004.

12. Choi E, Biswal S, Malin B, Duke J, Stewart WF, Sun J, editors. Generating multi-label discrete patient records using generative adversarial networks. Machine learning for healthcare conference; 2017: PMLR.

13. Kingma DP, Welling M. Auto-encoding variational bayes. arXiv preprint arXiv:13126114. 2013.

14. Lee D, Yu H, Jiang X, Rogith D, Gudala M, Tejani M, et al. Generating sequential electronic health records using dual adversarial autoencoder. Journal of the American Medical Informatics Association. 2020;27(9):1411–9.

15. Johnson AE, Pollard TJ, Shen L, Li-wei HL, Feng M, Ghassemi M, et al. MIMIC-III, a freely accessible critical care database. Scientific data. 2016;3:160035.

16. Muñoyerro-Muñiz D, Goicoechea-Salazar J, García-León F, Laguna-Tellez A, Larrocha-Mata D, Cardero-Rivas M. Health record linkage: Andalusian health population database. Gaceta Sanitaria. 2019;34(2):105–13.

17. Jordon J, Szpruch L, Houssiau F, Bottarelli M, Cherubin G, Maple C, et al. Synthetic Data--what, why and how? arXiv preprint arXiv:220503257. 2022.

18. Tong X, Yu Q, Ankawi G, Pang B, Yang B, Yang H. Insights into the role of renal biopsy in patients with T2DM: a literature review of global renal biopsy results. Diabetes Therapy. 2020;11(9):1983–99.

19. García-León F, Villegas-Portero R, Goicoechea-Salazar J, Muñoyerro-Muñiz D, Dopazo J. Impact assessment on data protection in research projects. Gaceta sanitaria. 2020;34(5):521–3.

20. Junta de Andalucía. Joint Resolution 1/2021 of the General Secretariat for Health Research, Development and Innovation of the Regional Ministry of Health and Families and the Management Directorate of the Andalusian Health Service. 2021 [Available from: https://www.sspa.juntadeandalucia.es/servicioandaluzdesalud/sites/default/files/sincfiles/wsas-media-sas_normativa_mediafile/2021/resolucion_conjunta_acceso_a_datos_investigacion_def_20211201%28F%29.pdf].

21. Muñoyerro-Muñiz D, Villegas-Portero R, de la Oliva V, Esteban-Medina A, Fernandez-Valle P, Sanchez A, et al. Ethical and Secure Evidence Generation from Regionwide Clinical Data through a Collaborative Environment for Advancing Predictive Care. Research Square. 2024;10.21203/rs.3.rs-5389651/v1.

22. European Commission. Regulation of the European Parliament and of the Council on the European Health Data Space 2024 [Available from: https://eur-lex.europa.eu/legal-content/EN/TXT/?uri=CELEX:52022PC0197].

23. Lou Y, Caruana R, Gehrke J, Hooker G, editors. Accurate intelligible models with pairwise interactions. Proceedings of the 19th ACM SIGKDD international conference on Knowledge discovery and data mining; 2013.

